# Burden of oral disorders in the U.S. from 1990 to 2023: Findings from GBD study

**DOI:** 10.64898/2026.01.18.26344340

**Authors:** Elnaz Tavazozadeh, Niki Shakour, Golsa Veysi, Kamyar Nasiri, Abdolreza Jamilian

## Abstract

**Objectives:** There is no comprehensive study comparing the burden of different oral diseases across states, years, and different demographics in the United States (US). Therefore, to address this gap, we aimed to evaluate the burden of oral disorders in the US and its states from 1990 to 2023.

**Methods:** We used the Global Burden of Disease (GBD) 2023 data to evaluate the burden of oral disorders in the USA from 1990 to 2023 in different age groups, sexes, states, and years. We aim to report the incidence, prevalence, and Disability-Adjusted Life Year (DALY) rates of caries of deciduous teeth, caries of permanent teeth, periodontal diseases, edentulism, and other oral disorders, accompanied by 95% uncertainty intervals (UIs).

**Results:** From 1990 to 2023, age-standardized prevalence, incidence, and DALY rates of total oral disorders in the USA declined slightly by 2.3%, 0.14%, and 4.82%, respectively. Among all oral disorders, caries of permanent teeth had the highest age-standardized prevalence and incidence rates, while edentulism accounted for the highest DALY burden in both 1990 and 2023. Males had higher incidence and prevalence, whereas females had higher DALY rates across all these 34 years. The burden of oral disorders varied by age and state, with older adults and states such as West Virginia showing the highest DALY rates.

**Conclusions:** Although the decline in the burden of oral disorders in the US over the past decades was a promising trend, the recent increases are concerning and warrant attention from policymakers.

## Introduction

Oral disorders encompass a wide range of conditions, with dental caries and periodontal diseases being among the most common conditions worldwide, affecting more than 3 billion people (1). These disorders are responsible for $298 billion of direct and $144 billion of indirect annual costs, which impose a considerable burden on healthcare systems, given that they account for more than 4% of total health-related costs (2). Although these disorders do not directly cause mortality, they adversely affect patients’ quality of life and contribute to a decline in quality-adjusted life expectancy, which is particularly important because most of these conditions are chronic and can have long-lasting effects on patients (3–5). Given the high prevalence of these disorders and their impact on overall quality of life, the World Health Organization (WHO) has made significant efforts over the past decades to increase knowledge and awareness about oral disorders and to support programs that promote oral health through preventive measures (6). Despite the efforts made to prevent oral diseases, it is shown that the age-standardized incidence rate of these disorders has increased slightly from 1990 to 2019, possibly due to lifestyle and dietary changes over the past years, which could be indicative of health systems’ failure due to the mainly preventive nature of these diseases (4, 7, 8).

Despite the overall increasing trend in the incidence of oral disorders, there have been considerable regional variations in this regard (8). In particular, the United States (US) showed one of the largest increases in the incidence of oral disorders between 1990 and 2019 compared to other countries, placing it among the top three countries with the greatest increases in oral disorder incidence (4). Several reasons might have been contributing to the growing burden of oral disorders among the US population. Changes in the demographic characteristics of the US population may be a major contributing factor, as the population is aging and the number of decayed, missing, and filled teeth (DMFT) tends to be higher among older adults compared to younger ones (9). Another particularly important factor may be changes in lifestyle and dietary habits among the US population, as sugar consumption has increased over recent years, potentially contributing to the rising incidence of oral diseases (10, 11). Several factors, including existing health disparities, make the prevention and management of oral diseases more challenging in the US. For instance, significant gender, racial, and ethnic disparities exist in access to oral healthcare, with men, Black individuals, and Hispanic populations being less likely to receive such services (12). Additionally, there are substantial disparities across states in both dental visit rates and the prevalence of oral diseases (12).

In brief, the US is among the countries that have experienced a considerable rise in the incidence of oral diseases due to several factors, such as changes in the population’s demographics and lifestyles. Furthermore, existing disparities highlight the need for comprehensive preventive programs and accessible services to improve the overall oral health in the community. Epidemiological studies would be helpful in this regard, considering that they identify the populations who are at a higher risk of oral diseases to guide the preventive and screening programs (13, 14). However, while some studies have studied the risk factors and epidemiology of oral diseases among the US population (9, 15–18), there is no comprehensive study comparing the burden of different oral diseases across US states, years, and different demographics. Therefore, to address this gap, we aimed to evaluate the burden of oral disorders in the US and its states from 1990 to 2023.

## Methods

### Source

In this study, we used the Global Burden of Disease (GBD) 2023 data from the Institute for Health Metrics and Evaluation (IHME) to evaluate the burden of oral disorders across the USA from 1990 to 2023. In general, IHME aims to assess the burden and epidemiology of 375 diseases and injuries, as well as 88 modifiable risk factors, across 204 countries and territories (19). The exact methodology for the GBD study has been previously discussed (19, 20). Since this study was a secondary analysis of the GBD database and did not directly include human subjects, it did not require ethics approval.

### Conditions and indices

We included caries of deciduous teeth, caries of permanent teeth, periodontal diseases, edentulism, and other oral disorders in our analysis while excluding all oral and lip cancers. We retrieved data on the incidence, prevalence, and DALY rates of oral disorders across age groups, sexes, states, and years. DALY is calculated by summing the years of healthy life lost due to disability and years of life lost from mortality, and indicates the total burden of a condition (21). All rates in this study are per 100,000 people for each index and are accompanied by 95% uncertainty intervals (UIs). In addition, we employed the socio-demographic index (SDI) from the GBD study, which represents a country’s development by taking per capita income distribution, the fertility rate of people under 25 years old, and mean years of education for adolescents and adults aged 15 and above into account. SDI ranges from 0 to 1, with greater values indicating better socio-demographic status (22).

### Statistical analysis

We used R version 4.5.2 for data analysis and visualization, using the following packages: cowplot (version 1.2.0), ggplot2 (version 4.0.1), tidyverse (version 2.0.0), and dplyr (version 1.1.4). We conducted locally estimated scatterplot smoothing (LOESS) regression to examine the predicted relationship between SDI and age-standardized DALY rates across the USA states from 1990 to 2023.

## Results

The age-standardized prevalence rate of total oral disorders decreased by 2.3% (95% UI: -5.88 to 1.69) from 37,407.66 per 100,000 people in 1990 to 36,547.72 per 100,000 people in 2023. Similarly, age-standardized incidence and DALY rates decreased. Oral disorders had an age-standardized incidence rate of 49,041.93 per 100,000 people in 1990, compared to 48,974.53 in 2023, representing a change of 0.14% (95% UI: -4.11 to 4.94). Age-standardized DALY rates for total oral disorders were 285.97 and 272.19 per 100,000 people in 1990 and 2023, respectively, representing a 4.82% decline (95% UI: -9.88 to 0.37). In both 1990 and 2023, caries of permanent teeth had the highest share of age-standardized prevalence (20,396.18 and 19,048.1 per 100,000 people, respectively) and incidence rates (32,268.18 and 30,732.31 per 100,000 people, respectively). In contrast, edentulism had the highest age-standardized DALY rates in both 1990 (148.34 per 100,000 people) and 2023 (140.5 per 100,000). Age-standardized prevalence, incidence, and DALY rates of each oral disorder decreased in 2023 compared to 1990, except for caries of deciduous teeth, which increased by 20.16% (95 UI: 7.56 to 36.82), 10.29% (95% UI: - 1.39 to 32.23), and 19.76% (95 UI: 6.64 to 40.07), respectively. Table 1 shows details of the changes in these measures.

**Table 1.**
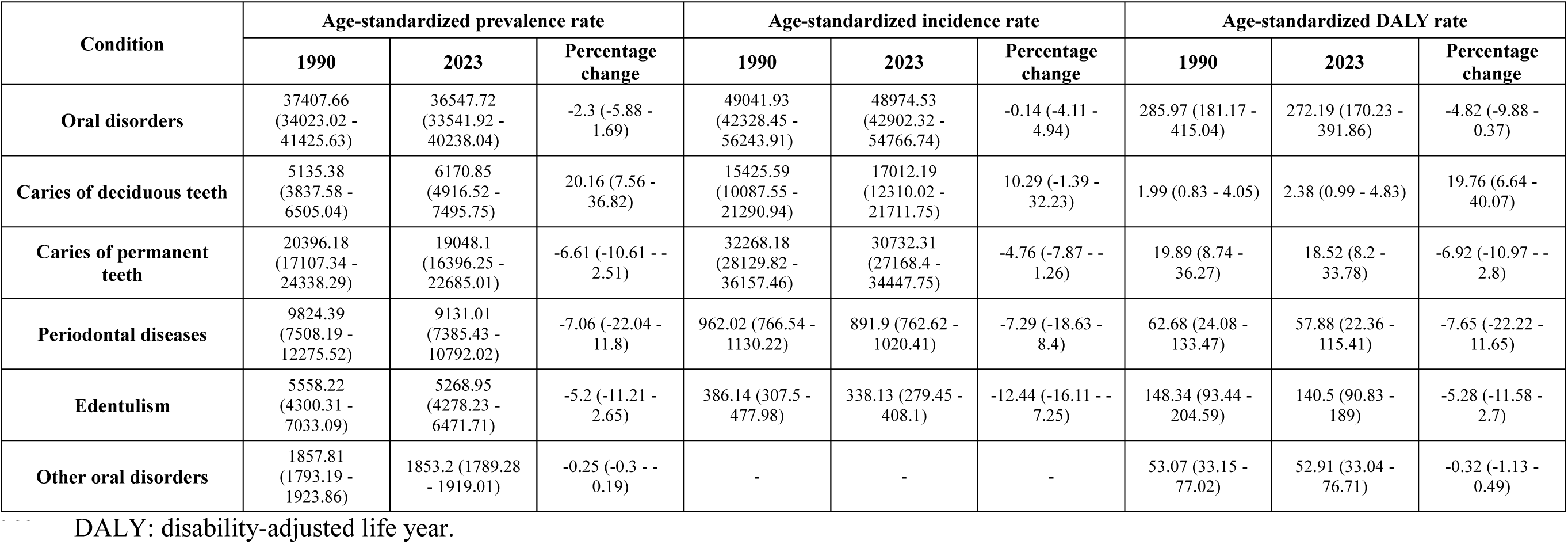
Burden of oral disorders in the United States in 1990 and 2021.

Age-standardized incidence, prevalence, and DALY rates of oral disorders in females and males across 1990 to 2023 are shown in Figure 1. Across all these years, the age-standardized incidence and prevalence rates of oral disorders in males were higher than those in females. On the contrary, females have consistently shown higher age-standardized DALY rates from 1990 to 2023. Figure 2 demonstrates incidence, prevalence, and DALY rates of oral disorders in males and females across all age groups. In both 1990 and 2023, the 5-9-year age group had the highest incidence rates of oral disorders in females (116,388 and 125868.1 per 100,00 people) and males (116,069.8 and 126,447.5 per 100,000 people). Moreover, the 80-84-year age group had the highest prevalence rates in females in 1990 (62,069 per 100,000 people) and 2023 (57,707.08 per 100,000 people). In 1990, males aged 85-89 had the highest prevalence of oral disorders (63,167.25 per 100,000 people), whereas in 2023, the 80-84-year age group had the highest rates (56,860.39 per 100,000 people). Finally, in females, DALY rates were highest in the 85-89- and 80-84-year age groups in 1990 and 2023 (1,121.72 and 999.01 per 100,000 people, respectively). Meanwhile, in both 1990 and 2023, the highest DALY rates among males were in the +95-year age group (1,284.88 and 980.15 per 100,000 people, respectively).

**Figure 1.**
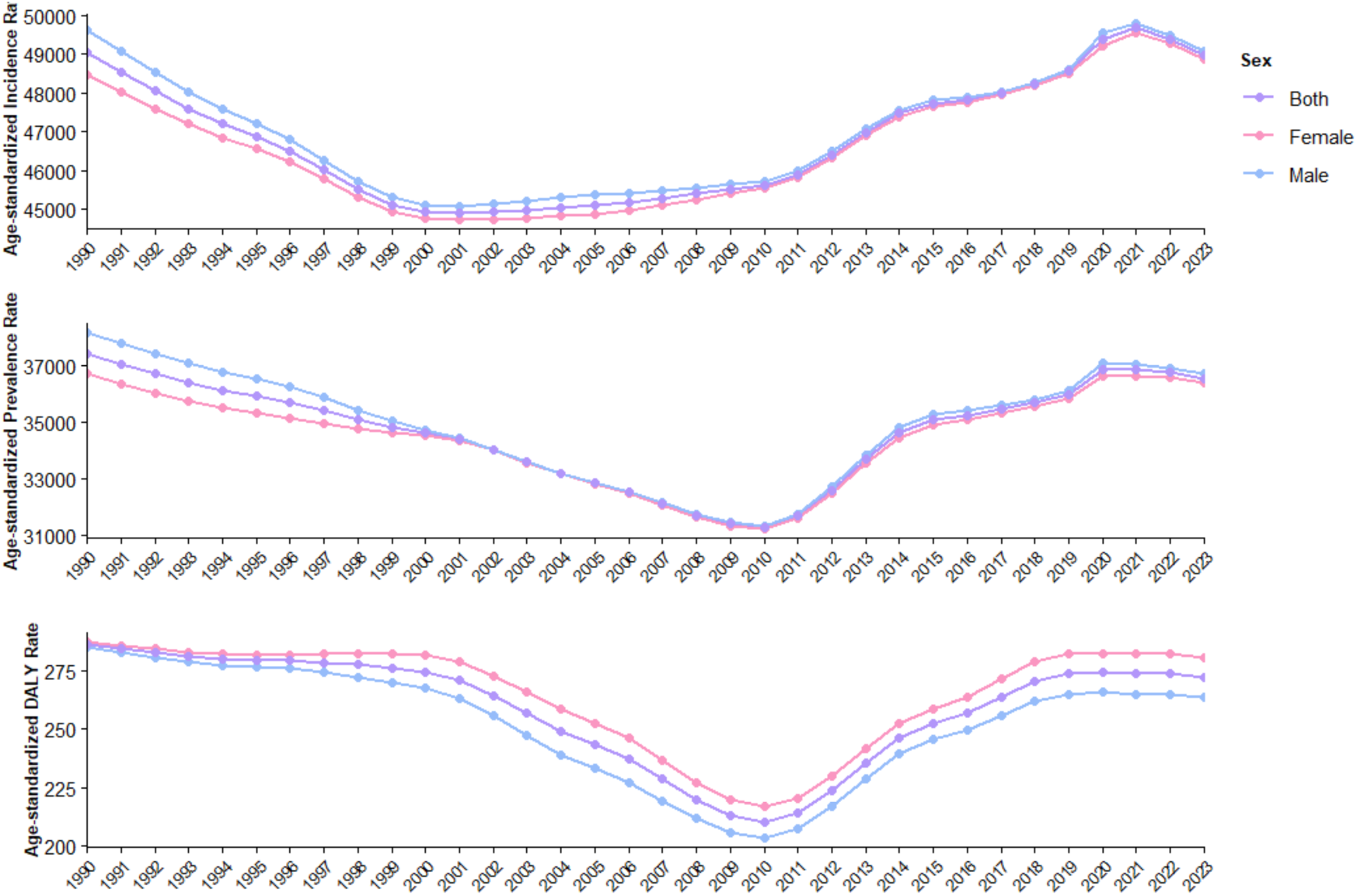
Trends in the burden of oral disorders across sexes from 1990 to 2023 in the US

**Figure 2.**
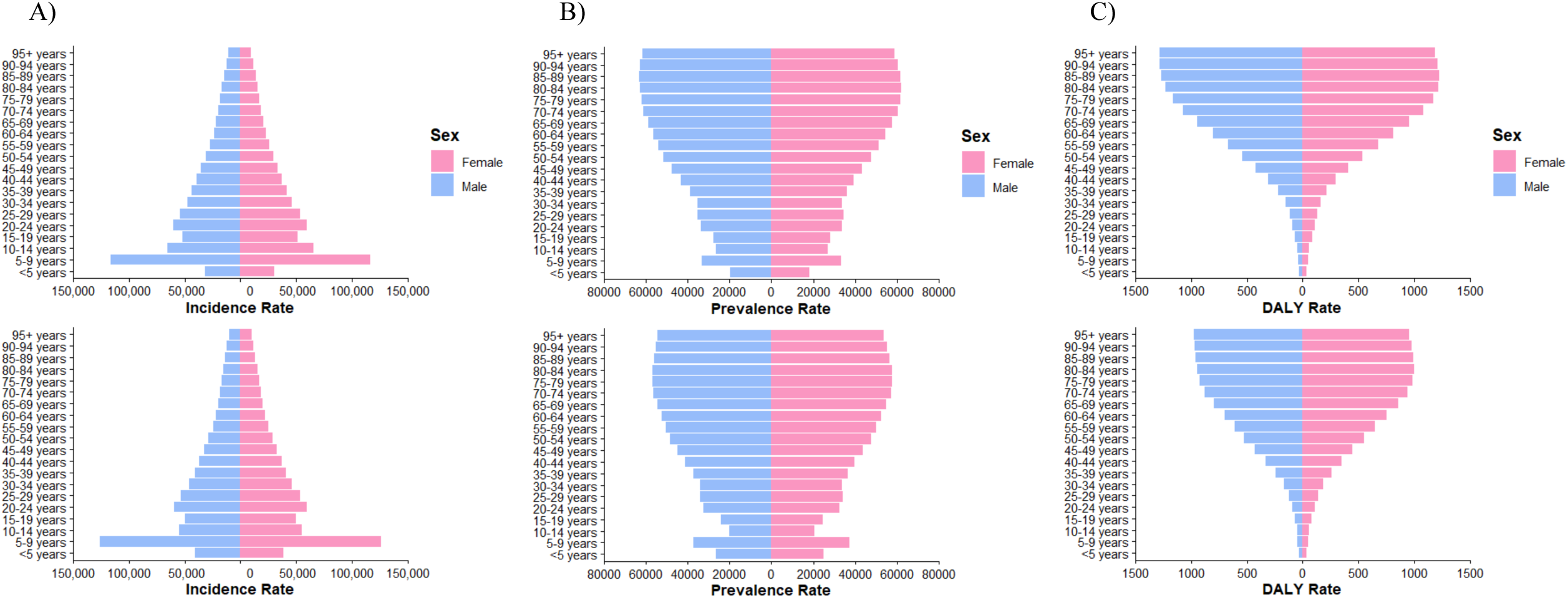
The incidence (A), prevalence (B), and DALY (C) rates of oral disorders across the sexes and age groups in 1990 (top) and 2023 (bottom) in the US

Figure 3 shows the incidence, prevalence, and DALY rates for each oral disorder across all age groups. In both 1990 and 2023, among people aged 10 and older, the incidence of caries of permanent teeth was higher than that of other oral disorders. Temporal changes in the prevalence rates of each oral disorder, however, were more complex. In 1990, among individuals younger than 10 years, caries of deciduous teeth showed the greatest prevalence. Among those aged 10 to 44 years, caries of permanent teeth had the highest prevalence. Adults aged 45 to 69 years were most affected by periodontal diseases, and in those aged 70 years and older, edentulism exhibited the highest prevalence rate. In 2023, the patterns of prevalence rates were quite similar, except for the 45-49-year age group, when caries of permanent teeth was the most prevalent oral disorder among people. Regarding the DALY rates, in 1990, edentulism had the highest DALY in people aged 45 and older, while in 2023, it had the highest DALY in people aged 35 and older.

**Figure 3.**
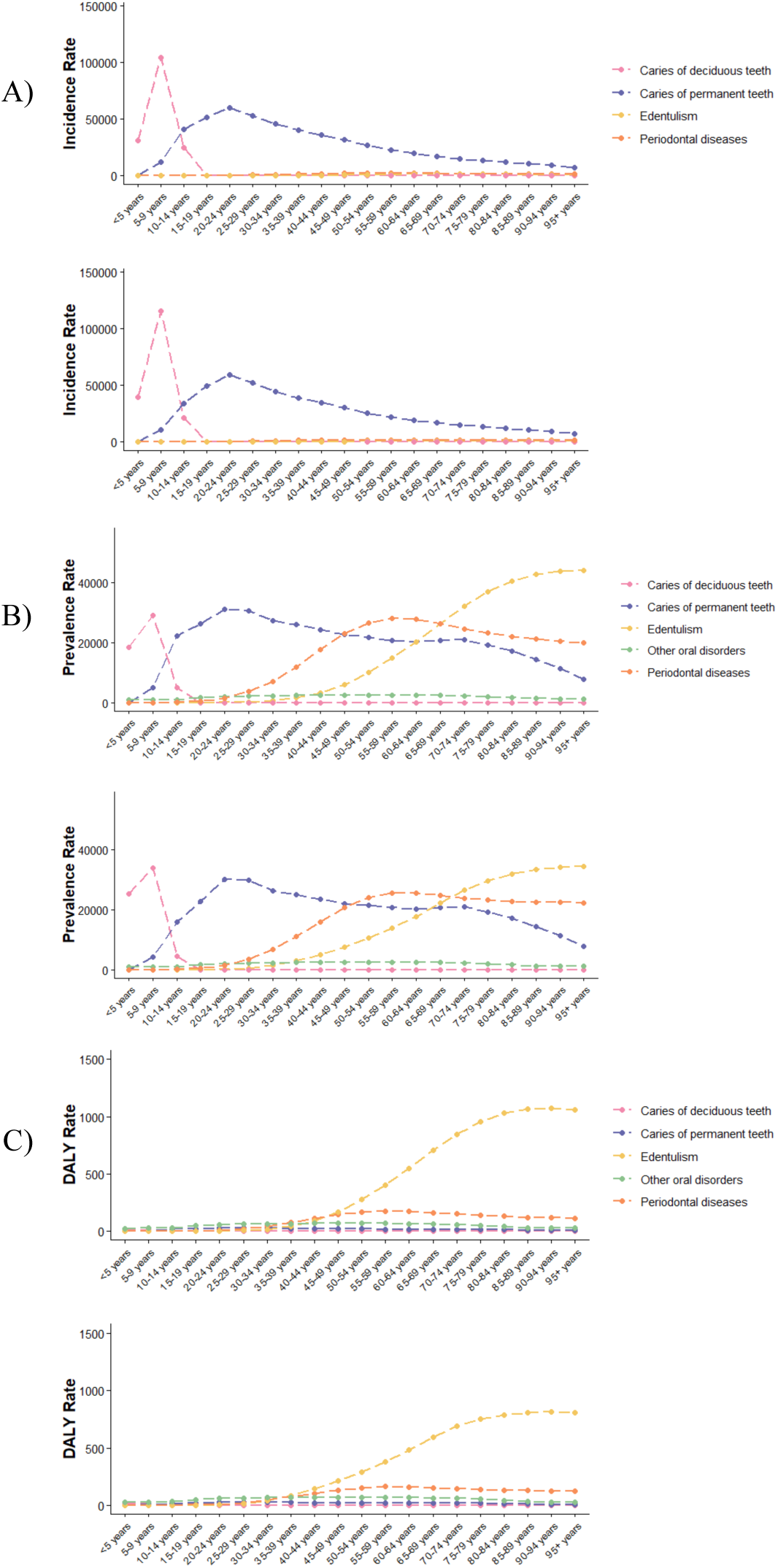
The incidence (A), prevalence (B), and DALY (C) rates of each oral disorder across the age groups in 1990 (top) and 2023 (bottom) in the US

As presented in Figure 4 and Figure 5, the age-standardized DALY rates of oral disorders were the highest in West Virginia (308.31 per 100,000 people), Tennessee (298.3 per 100,000 people), and Kentucky (297.70 per 100,000 people) in 1990, while West Virginia (308.7 per 100,000 people), North Carolina (303.77 per 100,000 people), and Tennessee (294.23 per 100,000 people) had the highest age-standardized DALY rates in 2023. In both 1990 and 2023, Nevada (276.51 and 258.46 per 100,000 people), Wisconsin (273.44 and 248.76 per 100,000 people), and Iowa (243.92 and 243.37 per 100,000 people) had the lowest age-standardized DALY rates of oral disorders. As evident in the figure, across all USA states, the age-standardized DALY rate for edentulism was higher than for other conditions.

**Figure 4.**
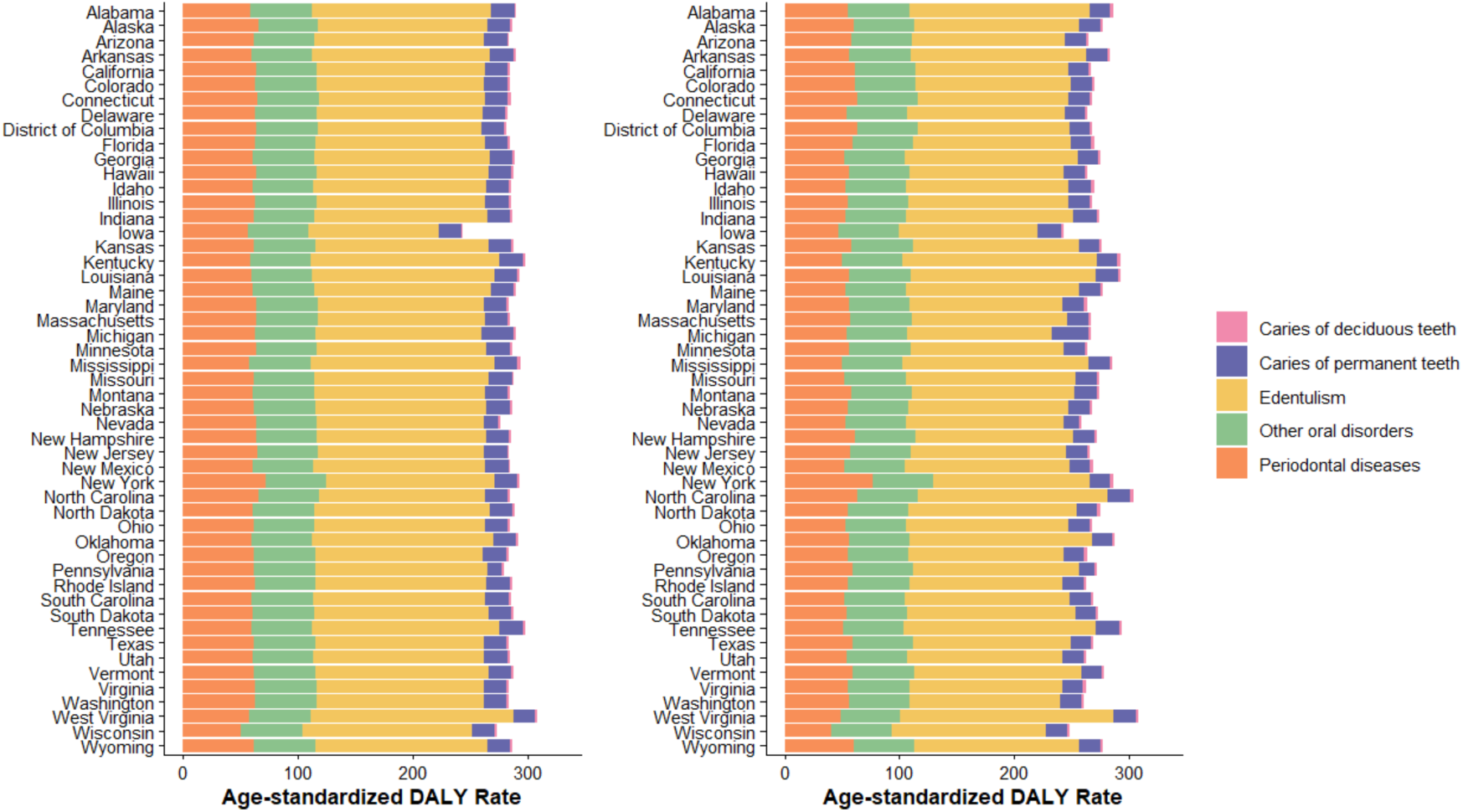
Age-standardized DALY rates of oral disorders in each US state in 1990 (left) and 2023 (right).

**Figure 5.**
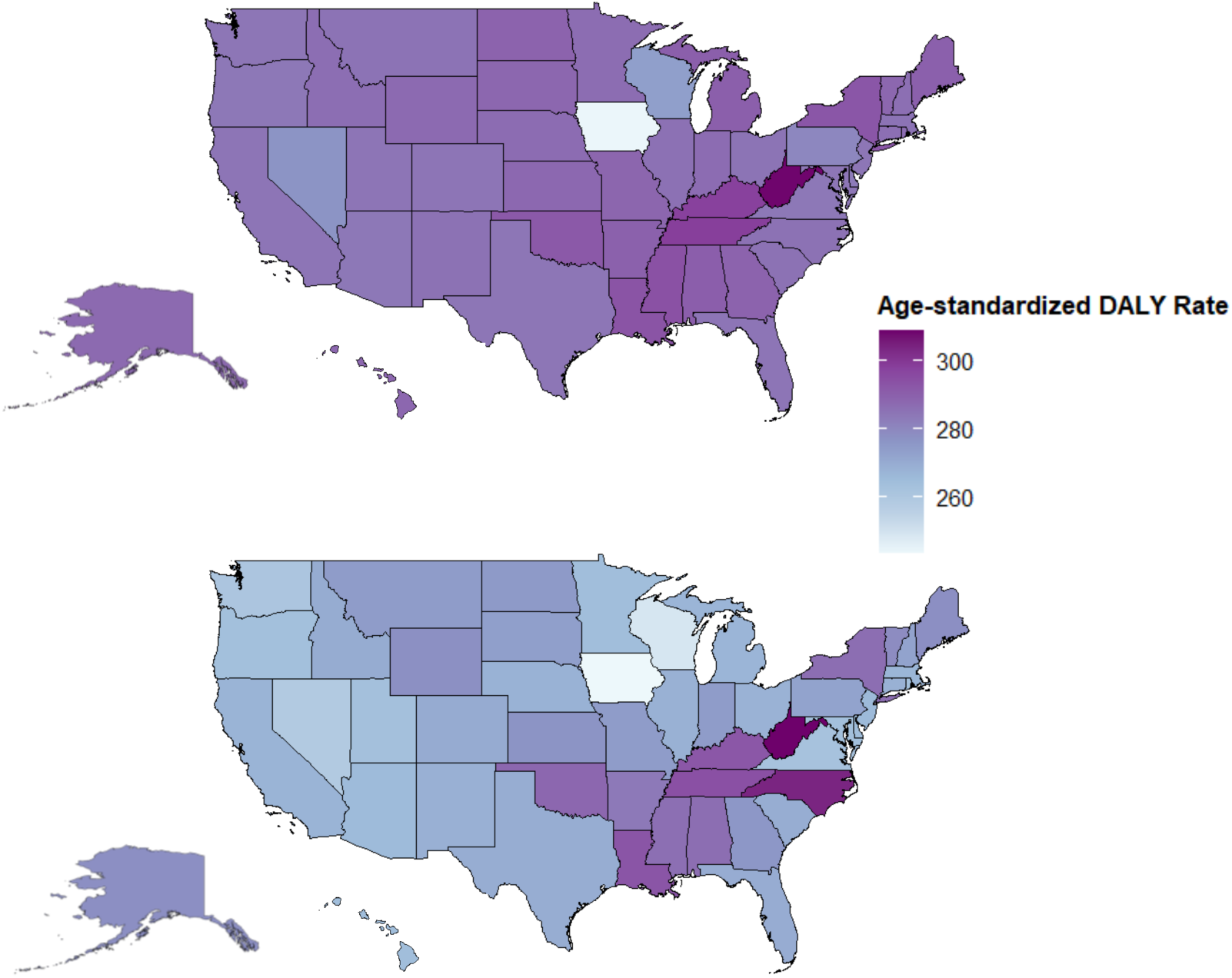
Choropleth map of age-standardized DALY rates of oral disorders in the US in 1990 (top) and 2023 (bottom).

The association between SDI and age-standardized DALY rates for oral disorders across time in the USA is shown in Figure 6. The expected value line had an ascending slope at lower SDIs (up to roughly 0.75), a descending slope at middle SDIs (roughly 0.75-0.85), and another ascending slope at higher SDIs (roughly more than 0.85). A descending slope indicates a negative association between SDI and age-standardized DALY rates, and an ascending slope suggests a positive association.

**Figure 6.**
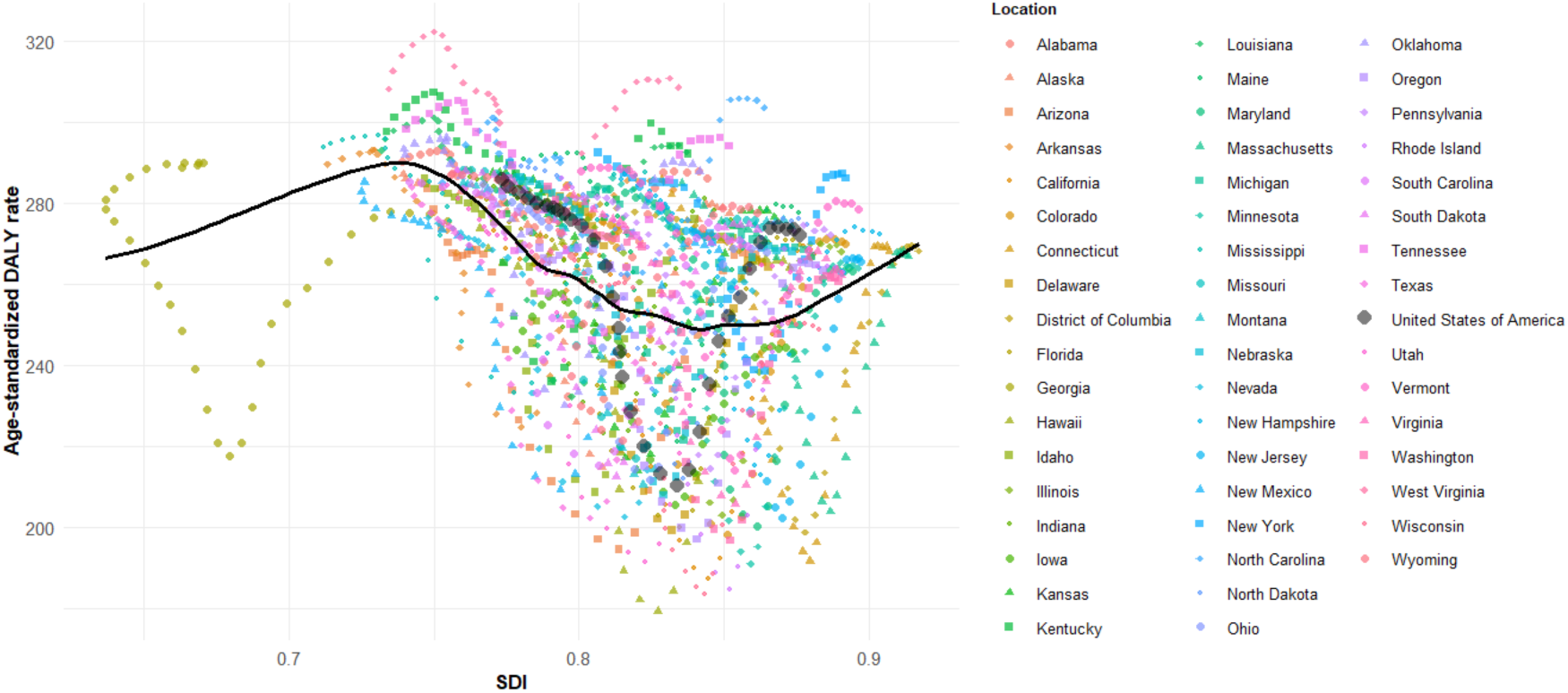
The association between the age-standardized DALY rates of oral disorders and sociodemographic indices in the US states

## Discussion

To the best of our knowledge, this study was the most comprehensive one on the epidemiology of oral disorders in the US, which could serve as a basis for future studies on these disorders and guide the preventive policies in the country. Throughout the study period, caries of permanent teeth had the highest burden compared to other disorders. Furthermore, while the burden of all diseases decreased over time, the burden of caries of deciduous teeth had an increasing trend. Nevada, Wisconsin, and Iowa had the lowest burden of oral disorders, while West Virginia and Tennessee remained among the top three states with the highest burden of these disorders throughout the study period. Additionally, while the age-standardized incidence and prevalence rates of oral disorders were higher among males, the age-standardized DALY rate remained higher in females from 1990 to 2023.

Even though the burden of oral disorders decreased between 1990 to 2023, the overall trend significantly differed over different periods. The age-standardized incidence rate of oral disorders decreased between 1990 and the early 2000s, and then gradually increased till 2023. Similarly, the age-standardized prevalence and DALY rate of oral disorders decreased till the late 2000s, and then started to increase. Therefore, the whole study period could be divided into two different parts. The overall decreasing pattern in the 1990s could have been due to several reasons. First, although water fluoridation in the US began in 1945, it was not until the early 1990s that more than 90% of the population had access to optimally fluoridated water, which might have had a crucial role in reducing the incidence of dental caries among US people, regardless of their socioeconomic status, in the subsequent years (23–26). Second, other preventive programs, such as the use of dental sealants, were implemented during that period and were well supported by policymakers and healthcare providers, which may have contributed to the declining burden of oral diseases at the time (27). However, these positive trends did not continue into the 2000s, and the subsequent increase in the burden of oral disorders may be attributed to several contributing factors. One possible explanation could be changes in the population’s demographics, such as an aging population and an increase in the proportion of Hispanic individuals, both of whom are at higher risk for oral diseases (9). These changes reflect a shift in the population’s needs, which may have gone unaddressed in recent years, as policies have failed to evolve accordingly to meet these emerging demands (28). Additionally, Medicaid’s limited coverage for dental care may have restricted access to oral health services for individuals from low socioeconomic backgrounds, contributing to disparities and a rising prevalence of oral diseases, particularly within this group (29, 30). Taken together, these findings suggest that the policies once effective in reducing the burden of oral disorders in the US have not been adequately refined to address the population’s evolving needs. As a result, the burden of oral diseases has begun to rise again and, if left unaddressed, may surpass the levels observed in 1990 in the coming years. Therefore, there should be a reassessment of the existing policies to modify them according to the unique needs of the US population in the coming years.

The burden of oral diseases remained the lowest in Nevada, Wisconsin, and Iowa throughout the study period, and these states may serve as role models for other states with regard to their policies for oral hygiene. A number of policies and initiatives in these states could have been responsible for the lower burden of oral disorders in these states. For example, in Iowa, about 90% of dental hygienists provide care to underserved populations and patients with special needs, and they are interested in supporting preventive programs in the community, which may play a crucial role in reducing oral health disparities, as these groups are at higher risk for dental diseases and have less access to dental care (31, 32). In Wisconsin, despite the existing disparities across people with different socioeconomic statuses, the oral health literacy was relatively high, which might be crucial in the lower burden of oral diseases in this state (33). Although further research is needed to identify the factors contributing to geographical disparities across US states, improving oral health literacy within communities and expanding care for underserved populations could be key strategies for reducing the national burden of oral diseases. The successful experiences of states such as Nevada, Wisconsin, and Iowa may offer valuable guidance for developing effective policies in other regions.

Even though this study is the most comprehensive one on the burden and epidemiology of oral disorders in the US, it has several limitations that are worth mentioning. First, although we identified populations with a higher burden of oral disorders, we did not assess the associated risk factors in our study. Therefore, we were unable to estimate the extent to which each specific factor, such as sugar consumption, contributed to the overall burden of oral disorders in the US. Future studies on the burden of oral disorders attributable to different risk factors across different demographic groups in the US could be beneficial in further guiding preventive policies. Second, we were unable to further explore the burden of oral disorders within ethnic and racial groups, where there have been found to be disparities with regard to oral health(12). Therefore, future studies could focus on examining racial and ethnic disparities in oral disorders and exploring how these patterns have evolved over time. Finally, the GBD study relies on estimations to determine the burden of diseases across different countries and regions, and the accuracy of the estimations depends on the quality of the primary data (34, 35). Although poor primary data quality is a more prominent issue in developing countries, future comprehensive studies in the US that utilize primary and registry data could help provide more accurate estimations at the national level.

### Conclusions

Although the decline in the burden of oral disorders in the US over the past decades was a promising trend, the recent increases are concerning and warrant attention from policymakers. Efforts to refine preventive strategies and improve treatment access are essential to counter this upward trend. States such as Nevada, Wisconsin, and Iowa have consistently shown lower burdens of oral disorders, and their successful policies may serve as valuable models for other states aiming to enhance overall oral health and reduce disease burden nationwide.

## Data Availability

All data produced in the present work are contained in the manuscript

## Ethical approval

This study utilized GBD data, which is publicly available. Given that no human subject was involved in the study, it didn’t require ethics approval.

## Author contributions

ET: Conceptualization, Methodology, Writing – review & editing

NS: Formal Analysis, Writing – original draft, Writing – review & editing

GV: Methodology, Writing – original draft, Writing – review & editing

KN: Writing – original draft, Writing – review & editing

AJ: Conceptualization, Writing – review & editing

